# Does working status affect the mental health outcomes of older population: A comparative study using the Longitudinal Ageing Study in India

**DOI:** 10.1101/2025.08.03.25332902

**Authors:** Poulomi Chowdhury, Mausam Kumar Garg, Amit Arora

## Abstract

Population aging is rapidly emerging as a global concern, with the proportion of older individuals rising steadily. In India, this demographic shift carries significant socioeconomic consequences, often pushing older people into financial insecurity and extending their participation in the workforce beyond retirement age. While studies from developed countries have shown a positive impact of continued engagement in later life on the mental health of older individuals, the present study aims to examine whether this relationship holds in the Indian context, where older individuals often continue working out of financial necessity rather than personal passion or motivation. Using nationally representative data from the Longitudinal Ageing Study in India, the study finds that working beyond retirement age is associated with a reduced risk of depression and poor cognitive functioning among older individuals. These findings are consistent across both logistic regression and propensity score matching analyses. The matched analysis shows that older working people have a 4.0 percentage point lower likelihood of poor cognitive functioning and a 3.5 percentage point lower likelihood of experiencing depression (both P < 0.001) compared to their matched non-working counterparts. This highlights the positive role of later-life engagement in supporting mental health. The study also emphasizes the need to create age-inclusive work environments, provide opportunities for skill development, and adapt job roles to suit physical capacities, enabling older people to remain productive and mentally engaged.

## Introduction

Population aging is increasingly becoming a global issue, with the share of older individuals rising rapidly. This demographic shift has significant socio-economic implications, particularly in labor force composition and market outcomes [1, 2]. According to a 2017 United Nations report, about 60 percent of the world’s older population lives in developing countries [3]. Developed countries, having experienced aging earlier, have enacted policies to encourage older population to remain in the workforce beyond the age of 65 [4–9]. However, in developing countries, pension systems and retirement programs are less widespread. In these nations, only around one-fifth of the older population receive pension benefits, leaving the majority dependent on family support [10]. Consequently, the proportion of older individuals in the workforce is higher compared to developed nations [11–16]. Therefore, it becomes essential to understand the relationship between employment and the quality of aging in-order to promote healthy and active. In addition, the discussion on work and the older population often centers on the concepts of productive versus unproductive aging [17–19] and active versus inactive aging [20]. In many developing countries, later life engagement is often a survival strategy, especially where a significant portion of employment is informal with poor social protection [1, 16, 21, 22].

The phenomenon of aging has become a widely discussed issue in India, with the older population projected to reach 319 million by 2050 [23]. This demographic shift is expected to surpass the younger population, leading to significant public health concerns [24], as aging is commonly associated with a higher risk of chronic health conditions [25–28]. With the rising burden of non-communicable diseases (NCDs), India is already experiencing an increasing demand for a subsidized public health system [29]. This trend is anticipated to escalate, placing a substantial financial strain on older people. Previous research has shown that between 39 to 50 million households in India fall into poverty due to high out-of-pocket healthcare payments [30, 31]. Despite this, most of the older population in India remains uncovered by any financial protection schemes [32–35].

Although several programs, such as the National Policy on Older Persons, National Policy for Senior Citizens, National Social Assistance Program, National Program for Healthcare of the Older People, Mahatma Gandhi National Rural Employment Guarantee Scheme, and Indira Gandhi National Old Age Pension Scheme, were introduced to provide social protection to older people in India, they have largely failed to achieve their goals due to issues related to coverage, implementation, corruption, and lack of awareness [36–39]. Additionally, the Indian government launched initiatives such as the Rashtriya Swasthya Bima Yojana (RSBY) and the Ayushman Bharat Program (ABP) to provide universal health coverage, especially targeting impoverished sections of society. However, these programs have faced significant challenges, including corruption, lack of awareness, and insufficient participation [32, 40]. As a result, to cope with financial insecurity and rising healthcare needs, older people often rely on their children or continue to work beyond retirement age [13, 16, 21, 41]. According to the 2011 Census of India, approximately 33 million older people are working beyond the retirement age [42]. Studies in developing countries have highlighted that factor such as gender [11, 12, 14, 16, 19, 43, 44], occupation type [11–13, 16, 21, 43], education [11, 16, 19, 43], and place of residence [11, 16, 43, 45, 46] are significant determinants of later-life work engagement. Moreover, working in later life is closely linked to mental health [9, 12, 47–51]. Although few international research has examined the effects of working later in life on mental health, there is a no national studies on this topic. As India faces an aging population, promoting healthy and active aging will become increasingly important. Therefore, present study tries to answer an important research question “does working status (working/non-working) affects the mental health outcomes of the older population?”. Research findings will help provide an insight to the policymakers and future research about the potential benefits of active and productive ageing on the mental health.

## Materials and methods

### Data source

This paper draws on data from the first wave of the Longitudinal Ageing Study in India (LASI), a cross-sectional survey conducted in 2017-18 [52]. It is a nationally representative survey, led by the International Institute for Population Sciences (IIPS) in collaboration with the Harvard T.H. Chan School of Public Health and the University of Southern California, collected data from 72,250 Indian adults aged 45 and above. Using a multistage-stratified cluster sampling design, the survey gathered information on various aspects such as social and economic well-being, mental health, functional health, chronic health conditions, healthcare utilization and costs, and childhood health experiences [52]. From the total sample, 31,464 older individuals aged 60 and above were selected for this study. Among them, approximately 10,746 are working and 20,718 are non-working older population.

### Outcome variables

In line with the research question, this current study focuses on two mental health outcomes: poor cognitive functioning (PCF) and depression. The details of these outcomes are outlined below:

#### Poor Cognitive Functioning

To assess poor cognitive functioning, this study follows the LASI report, which utilizes the cognitive module from the Health and Retirement Study (HRS). This module includes domains such as memory (0–20), orientation (0–8), retrieval fluency (0–61), arithmetic function (0–9), executive function (0–4), and object naming (0–2). The first step in the analysis is normalizing these indicators using the following formula:

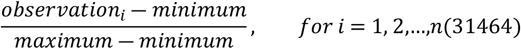

This normalization process rescales the indicators to a range between 0 and 1 [53]. Following this, a principal component analysis is conducted to generate a composite cognitive functioning score, where a lower score indicates poorer cognitive functioning, and a higher score represents better cognitive performance [54, 55]. The composite score is then divided into three equal tertiles, with the first tertile coded as 1 (indicating poor cognitive functioning) and the remaining tertiles coded as 0 (otherwise) [13].

#### Depression

It is assessed using the self-reported Centre for Epidemiological Studies Depression (CES-D) scale, which categorizes responses into four groups: rarely or never (<1 day), sometimes (1–2 days), often (3–4 days), and most or all of the time (5–7 days). Participants answered ten questions about their experiences in the week leading up to the interview. Of the 10 items, the first seven measure negative symptoms, while the remaining three measure positive symptoms. For negative symptoms, a score of zero is assigned to responses of “rarely or never (<1 day)” or “sometimes (1–2 days),” with the other two categories scored as one. Scoring for positive symptoms is reversed. The total score ranges from 0 to 10. Participants scoring four or higher are considered to have depressive symptoms [56, 57] and are coded as 1, while those scoring below four are coded as 0 [1, 41, 52, 58].

### Exposure Variable: Working status

The main exposure variable in this study is working status of the older population, categorized as 1 for working and 0 for non-working.

### Other covariates

The selection of potential covariates is based on prior research related to the relationship between working status and mental health of the older population [9, 12, 13, 47–51]. These factors are grouped into three key dimensions: socio-economic and demographic characteristics, health condition, and lifestyle behaviours. The socio-economic and demographic dimension consists of gender (male, female), place of residence (rural, urban), education level (low, middle, high), wealth index (low, medium, high), caste groups (general, scheduled tribe (ST), scheduled caste (SC), other backward class (OBC)), and religion (Hindu, Muslim, Others). Further, health condition includes self-rated health (good/fair, poor), while lifestyle behaviours consist of smoking/ consuming tobacco (no, yes), drinking alcohol (no, yes), and yoga (no, yes).

### Statistical analysis

Bivariate and multivariate analyses were employed for statistical analysis. In the bivariate analysis, the percentage of working status was calculated across various background variables, and the prevalence of PCF and depression was estimated based on working status. A chi-square test was conducted to assess the statistical significance of the associations.

For multivariate analysis, multiple sequential logistic regression was applied, with PCF and depression as the outcome variables. The final model was adjusted for all covariates. Although logistic regression gives us the true association between working status and mental health outcomes, however causality of this relationship cannot be established because of cross-sectional as well as non-experimental nature of the data. Therefore, propensity score matching (PSM) approach is utilized to overcome this shortcoming. This method has been increasingly adopted to reduce or eliminate the effects of confounding when using observational data [59]. Li (2012) highlights by adjusting the covariates between treated and control groups and reconstructing counterfactual outcomes, PSM enables the estimation of an unbiased causal effect using observational data.

#### Overview of PSM

Propensity score matching is a statistical technique designed to estimate treatment effects using non-experimental or observational data [60, 61]. The effect of a treatment on an individual, can be estimated as follows:

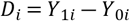

Where, *Di* denotes difference between mental health outcomes of *i*^*th*^ individual. While *Y*_1_ and *Y*_0_ represent the mental health outcomes for *i*^*th*^ working and non-working older population. However, *Y*_1*i*_ and *Y*_0*i*_ cannot be observed simultaneously, as the same unit cannot be part of both the treatment and control groups at the same time. The outcome that remains unobserved is referred to as the “counterfactual outcome” [62].

The average treatment effect (ATE) can be estimated as:

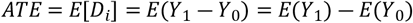

Where *E*(*Y*_1_) and *E*(*Y*_0_) represent the expected values of Y for all units in the treatment and control groups, respectively.

In observational studies, the key parameter of interest is often the ‘average treatment effect on the treated’ (ATT), which is calculated as:

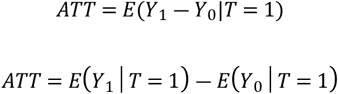

In this context, *T* = (0, 1) represents control and treatment conditions. The counterfactual mean, *E*(*Y*_0_| *T* = 1), is unobservable and must be substituted to estimate the ATT. If treatment assignment is random, *E*(*Y*_0_| *T* = 0) can be used as the substitute. However, in observational studies, direct comparisons are often misleading due to systematic differences between the treated and control groups [63].

Since observational data lacks randomized treatment assignment, statistical methods are needed to balance the data before assessing treatment effects [61]. PSM helps reduce bias by creating a balanced sample where confounding factors are controlled [64]. Propensity scores align the distribution of observed baseline covariates between treated and untreated subjects, mitigating selection bias in the matched sample [59, 61]. Further, all the analysis has been performed in STATA software.

## Results

Given Table 1 represents the percentage distribution of background characteristics. Among the total older population, approximately 48.4 percent of male and 38.7 percent of the rural population remain engaged in the workforce beyond retirement age. In contrast, about 79 percent of female belong to the non-working category. In addition, the proportion of older working population decreases with the increase in education levels and wealth index. The percentage of older workers is higher among STs and those with unhealthy lifestyle behaviours, such as drinking alcohol. Furthermore, roughly 73.8 percent of the older individuals with poor self-rated health fall into the non-working category.

**Table 1.** Background characteristics of older population (60 years and above)

| Covariates | Working Status |  | Number of samples |
| --- | --- | --- | --- |
|  | Non-Working | Working |  |
| **Gender** |  |  |  |
| Male | 51.6 | 48.4 | 15097 |
| Female | 79.0 | 21.0 | 16367 |
| **Place of residence** |  |  |  |
| Rural | 61.3 | 38.7 | 20726 |
| Urban | 74.7 | 25.3 | 10741 |
| **Education level** |  |  |  |
| Low | 65.5 | 34.5 | 24448 |
| Middle | 64.3 | 35.7 | 4761 |
| High | 73.2 | 26.8 | 2258 |
| **Wealth Index** |  |  |  |
| Low | 60.5 | 39.5 | 10555 |
| Medium | 64.2 | 35.8 | 10421 |
| High | 72.9 | 27.1 | 10491 |
| **Caste groups** |  |  |  |
| General | 74.7 | 25.3 | 9269 |
| Scheduled Tribe (ST) | 57.8 | 42.2 | 5172 |
| Scheduled Caste (SC) | 62.6 | 37.4 | 5140 |
| Other Backward Class (OBC) | 63.8 | 36.2 | 11886 |
| **Religion** |  |  |  |
| Hindu | 64.9 | 35.1 | 23035 |
| Muslim | 72.2 | 27.8 | 3737 |
| Others | 65.3 | 34.7 | 4695 |
| **Self-rated Health: Poor** | 73.8 | 26.2 | 7113 |
| **Drinking Alcohol: Yes** | 42.2 | 57.8 | 2883 |
| **Smoking/Consuming Tobacco: Yes** | 55.9 | 44.1 | 12176 |
| **Yoga: Yes** | 69.6 | 30.4 | 3416 |
| **Total sample** | 20718 | 10746 | 31464 |

### Working status and mental health outcomes

Fig-1 illustrates the prevalence of PCF and depression among working and non-working older population. The risk of PCF is significantly higher in the non-working older population compared to their working counterparts (P<0.001). Likewise, 29.4 percent of the non-working older population are more likely to experience depressive symptoms compared to 25.3 percent of the working older population (P<0.05).

**Fig 1.**
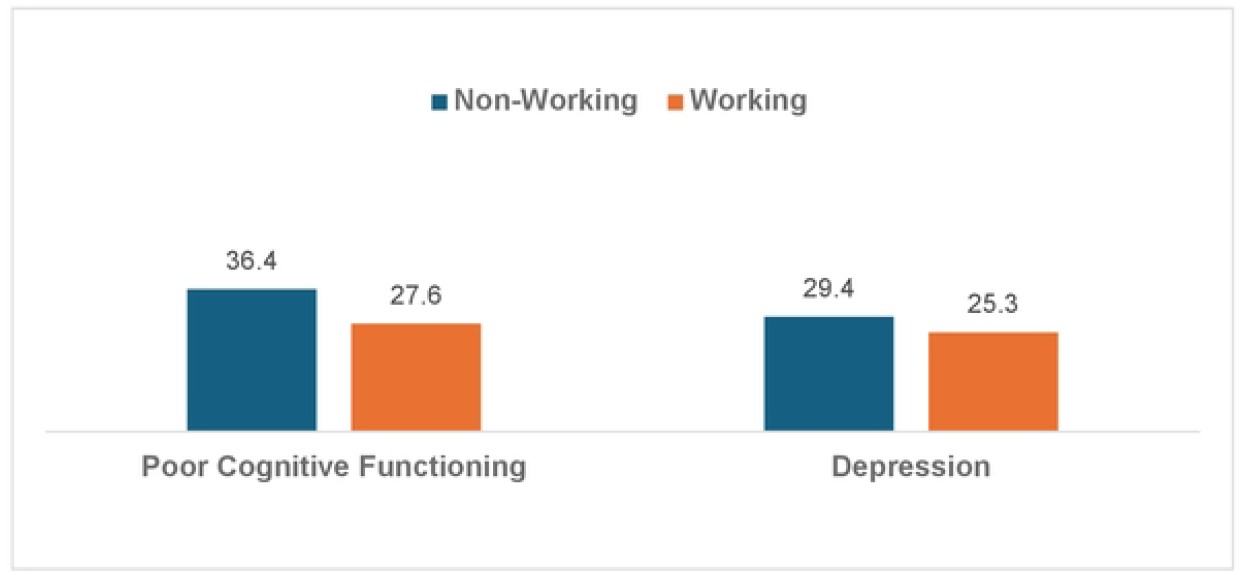
Prevalence of PCF and Depression by working status

Table 2 exhibits the logistic regression results for PCF and depression. Both the unadjusted and adjusted odds ratios indicate a significant association between working status and mental health outcomes. Older individuals working beyond the retirement age have 0.652 times (P<0.001) lower odds of PCF compared to their non-working counterparts. Likewise, the odds of depression are 0.849 times (P<0.001) lower among the working older population than that of non-working older population.

**Table 2.**
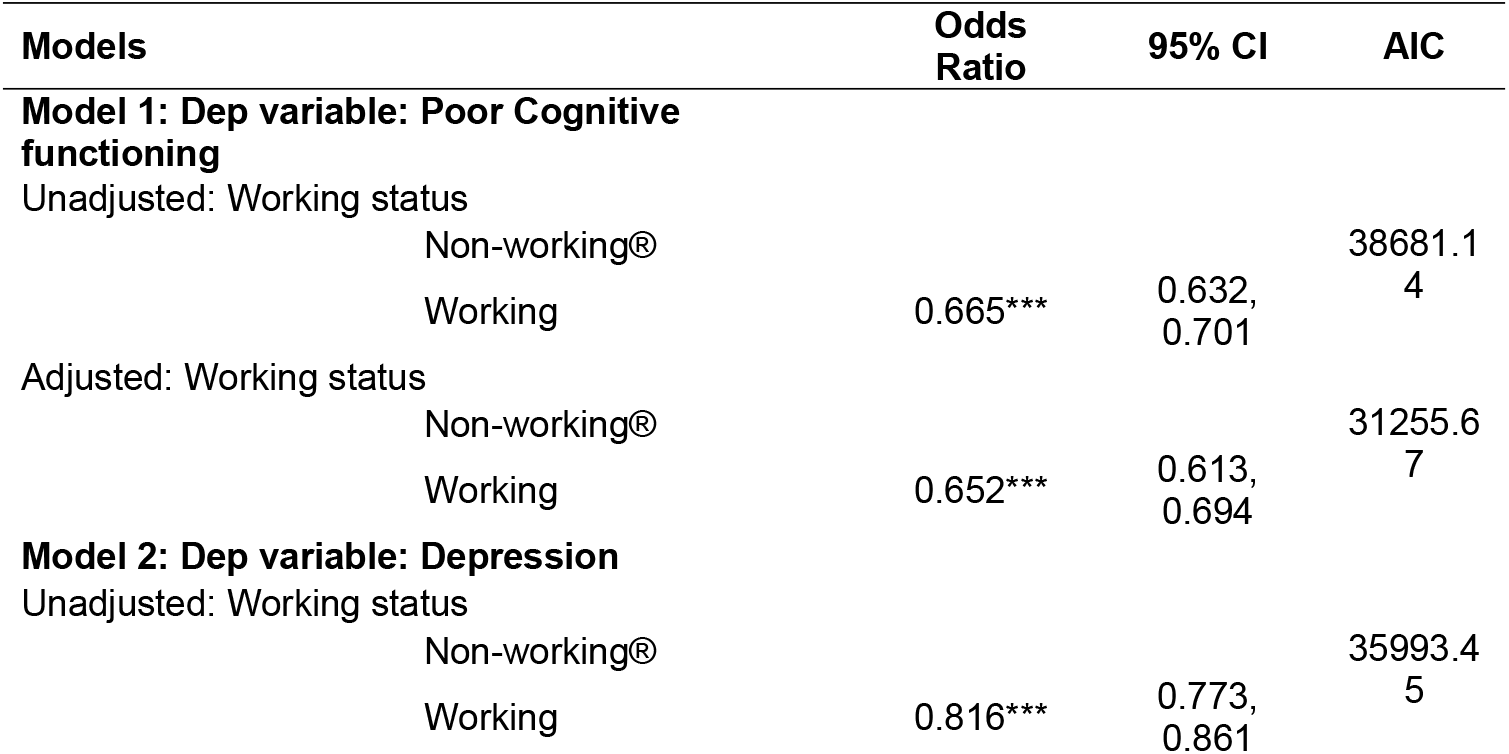
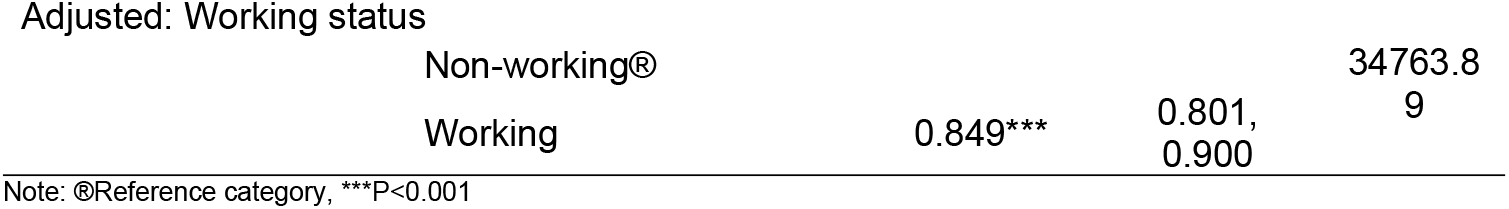
Result of logistic regression for PCF and Depression by working status.

### Estimates of PSM

Table 3 depicts the matching estimates from the propensity score analysis for PCF and depression, respectively. The estimates for ATE and ATET indicate that working status significantly affects both PCF and depression. The ATE for PCF is −0.037 (P<0.001), suggesting that working beyond retirement age reduces the risk of PCF by 3.7 percentage points. Similarly, the ATET shows that the propensity for PCF is 4 percentage points lower (P<0.001) among the working older population when matched with control groups.

**Table 3.** Matching estimates of PSM for PCF and depression.

| Non-Working vs Working | Coefficient | Robust SE |
| --- | --- | --- |
| **PCF** |  |  |
| ATE | -0.0366*** | 0.0069 |
| ATET | -0.0402*** | 0.0063 |
| **Depression** |  |  |
| ATE | -0.0252*** | 0.0070 |
| ATET | -0.0356*** | 0.0069 |

Note: \*\*\*P<0.001

For depression, the ATE of −0.025 (P<0.001) indicates a 2.5 percentage point reduction in the risk of depression among the older working population. The ATET value of −3.5 percentage points (P<0.001) further confirms that working beyond retirement age reduces the risk of depression when matched with control groups.

Fig 2 displays the kernel density plots of covariates across treatment levels for both the raw and matched samples. The plots are shown for both PCF and depression. If the kernel density plots for the matched sample are identical across treatment levels, it indicates that the covariates are balanced [65]. The density plots for the matched sample overlap perfectly (Fig 2), demonstrating that matching on the estimated propensity scores successfully balanced the covariates.

**Fig 2.**
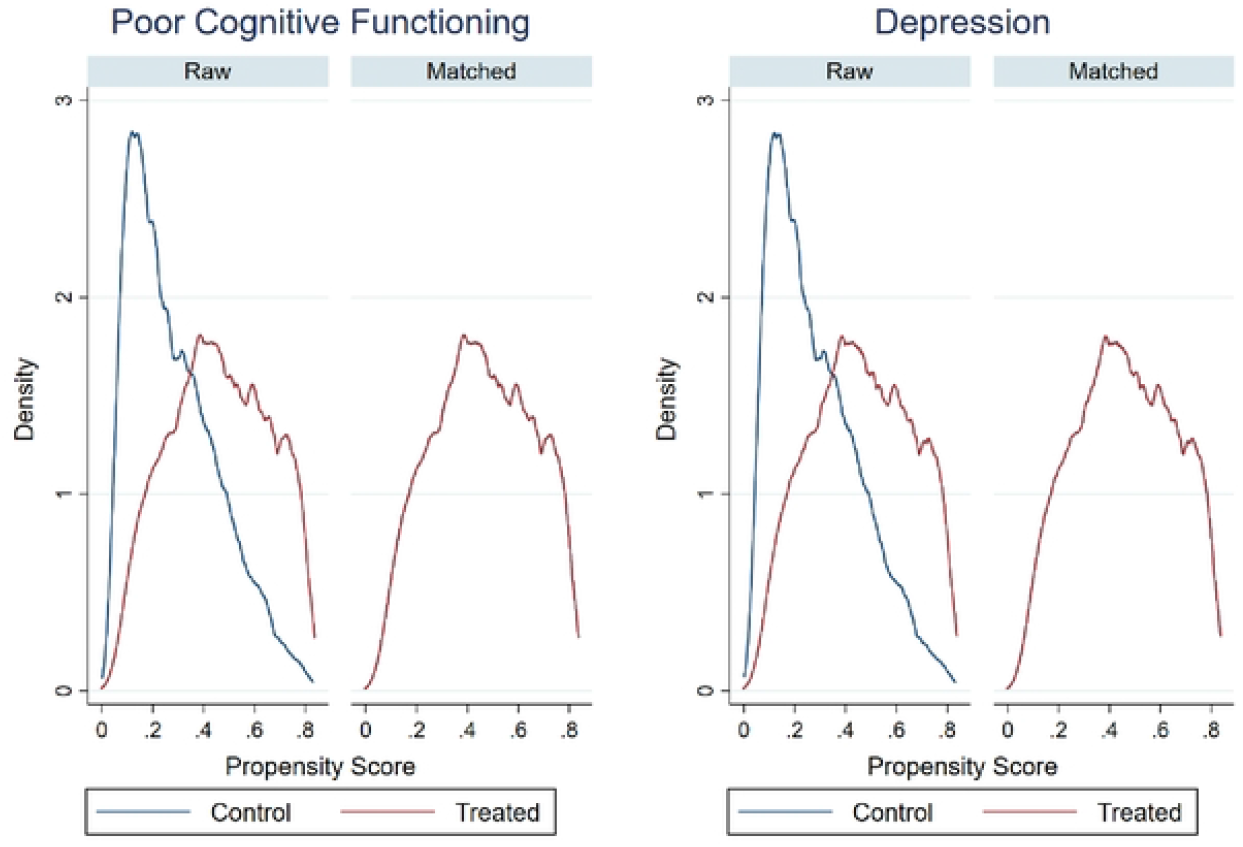
Kernal density plot

## Discussion

Globally, clinicians and institutions working with older population have expressed increasing concern about the mental health challenges associated with ageing. Further, growing research is investigating the determinants of mental health and exploring potential pathways to improve psychological well-being in later life. Mental health is now widely recognized as a crucial component of public health among older people, alongside physical health, due to its significant impact on overall well-being. While factors such as social connections, lifestyle behaviours, and voluntary activities have been widely studied, limited attention has been given to the role of working status in later life, particularly its association with cognitive functioning and depression. In many developing countries, older people often continue working beyond the official retirement age, primarily due to financial necessity. In India, where formal pension systems, healthcare benefits, and social protection schemes remain limited, extended workforce participation among older individuals is common. Against this backdrop, the present study aims to examine whether working status (working/non-working) in later life is associated with mental health outcomes among older people in India.

The findings of this study suggest that the prevalence of PCF and depression is significantly lower among older individuals who continue working beyond the retirement age compared to their non-working counterparts. This pattern is consistent with the results of logistic regression analysis, which indicates a strong positive association between later-life employment and improved mental health outcomes, even after adjusting for confounding variables. However, despite controlling for observable factors, logistic regression may still suffer from residual confounding due to unmeasured covariates, including potential interactions or non-linear effects.

To address this limitation and enhance causal inference, the study applied Propensity Score Matching. PSM creates a balanced comparison by matching working and non-working older people with similar observable characteristics. This approach reduces selection bias arising from the non-random nature of workforce participation, where those who continue working may differ systematically in health status, education, or socio-economic conditions. The matched analysis revealed that working older individuals had a 4.0 percentage point lower likelihood of PCF and a 3.5 percentage point lower likelihood of depression (both P < 0.001) compared to their matched counterparts. These negative and statistically significant ATET estimates suggest that employment beyond retirement age may exert a protective effect against mental health challenges in later life.

These findings align with evidence from international studies. For instance, Tomioka, Kurumatani (2018) found that continued employment among older men had a preventive effect against cognitive decline, although no significant relationship was observed for older women. Similarly, Li, Xu (2014) reported that older people engaged in productive activities experienced lower levels of depression, better functional and self-rated health. A systematic review by Baxter, Blank (2021) concluded that later-life employment often yields beneficial or neutral mental health outcomes, particularly among men and part-time workers. Silver, Dass (2018) also found that post-retirement employment reduced depressive symptoms in both men and women.

Numerous theoretical frameworks suggest that work provides a meaningful context that shapes cognitive functioning in later life [68]. Consistent with these perspectives, the present study underscores the potential benefits of continued work engagement on cognitive outcomes among older people. Empirical evidence also supports this association; for instance, Chowdhury, Mohanty (2023), Luo, Pan (2019) found that prolonged workforce participation is positively linked to cognitive health in the ageing population. Moreover, Fisher, Chaffee (2017) emphasizes the bidirectional relationship, noting that work can influence cognitive functioning, while cognitive abilities, in turn, affect work-related factors such as motivation, learning capacity, skill development, training outcomes, and safety.

Several factors may explain the mental health benefits of working beyond retirement age, especially in the Indian context. First, retirement can only be mentally and financially beneficial if individuals are covered by health insurance or social protection schemes. However, in India, only a small proportion of older people have access to such benefits [1, 69, 70]. Even universal schemes like Pradhan Mantri Jan Arogya Yojana (PMJAY) and RSBY are often marred by poor service delivery [32, 71–75]. As a result, retirement may exacerbate financial vulnerability due to the lack of income and the burden of out-of-pocket healthcare costs. Continued employment thus becomes a crucial financial safeguard, and financial security has been consistently linked to better mental health [47, 50].

Second, work provides older individuals with a sense of purpose and pride, contributing to household income and maintaining their role within the family. In addition, the workplace offers opportunities for social engagement, a factor strongly associated with improved mental well-being. Numerous studies underscore the importance of social connection in promoting mental health, particularly in later life [76–78].

This study has certain limitations, primarily the use of cross-sectional data collected at a single point in time. To gain a more comprehensive understanding of the relationship between working status and health, longitudinal data would be more suitable. However, in the Indian context, there is currently no longitudinal dataset that specifically captures the working status of older people. Despite this limitation, the study addresses potential bias by employing propensity score matching techniques, which enhance the robustness of the findings. Future research with a specific focus on the working older population, supported by a sufficiently large sample size, could offer stronger and more conclusive evidence of this relationship.

## Conclusion

The study highlights a strong link between working beyond retirement age and better mental health outcomes among older individuals. Those who continue working after the retirement age (60 years) show significantly lower risks of poor cognitive functioning and depression compared to their non-working counterparts. These findings are supported by both logistic regression and PSM analyses, indicating that continued engagement in work may have a protective effect on mental and cognitive well-being in later life. Thus, it is important to promote active ageing by encouraging older people to participate in flexible work suited to their capabilities. For example, participation of the 70 plus population in part-time or less physically demanding jobs can further support this goal and promote healthy and active ageing. In addition, creating age-inclusive work environments, offering opportunities to upgrade skills, and adapting roles to accommodate physical limitations can help older people remain productive and mentally engaged. All these can be achieved through strengthening pension, financial and social protection systems ensuring that older people are not compelled to work due to economic insecurity. A strong social protection scheme will allow older individuals to work only when they are motivated and passionate, ensuring dignity, autonomy, and improved quality of life. By implementing these policies, governments and organizations can help promote healthy and productive ageing, improving both the quality of life and mental health of the older population.

## Data Availability

This study is based on secondary data from the Longitudinal Ageing Study in India (LASI), which is publicly available upon request through the official IIPS website: https://www.iipsindia.ac.in/content/lasi-wave-i The authors confirm that they did not have any special access privileges and that others can obtain the data by following the same procedure.

## Acknowledgments

I sincerely thank IIPS for providing access to the Longitudinal Ageing Study in India dataset for this research.

## References

1. Chowdhury P, Goli S. Informal employment and high burden of out-of-pocket healthcare payments among older workers: evidence from the Longitudinal Ageing Study in India. Health Policy and Planning. 2025;40(2):123–39. doi: 10.1093/heapol/czae074.

2. Christensen K, Doblhammer G, Rau R, Vaupel JW. Ageing populations: the challenges ahead. The Lancet. 2009;374(9696):1196–208. doi: 10.1016/S0140-6736(09)61460-4.

3. UNITED NATIONS. World Population Ageing 2017-Highlights. Department of Economic and Social Affairs. 2017.

4. McAllister A, Bentley L, Bronnum-Hansen H, Jensen NK, Nylen L, Andersen I, et al. Inequalities in employment rates among older men and women in Canada, Denmark, Sweden and the UK. BMC Public Health. 2019;19(1):319. Epub 2019/03/20. doi: 10.1186/s12889-019-6594-7. PubMed PMID: 30885164; PubMed Central PMCID: PMCPMC6423867.

5. Larsen M, Pedersen PJ. Labour force activity after 65: what explain recent trends in Denmark, Germany and Sweden? J Labour Mark Res. 2017;50(1):15–27. Epub 2017/10/27. doi: 10.1007/s12651-017-0223-7. PubMed PMID: 29071306; PubMed Central PMCID: PMCPMC5632993.

6. Nilsson K, Ostergren PO, Kadefors R, Albin M. Has the participation of older employees in the workforce increased? Study of the total Swedish population regarding exit from working life. Scand J Public Health. 2016;44(5):506–16. Epub 2016/03/19. doi: 10.1177/1403494816637262. PubMed PMID: 26988576; PubMed Central PMCID: PMCPMC4887820.

7. Garcia MTM, Fontainha E, Passos J. Hiring older workers: The case of Portugal. The Journal of the Economics of Ageing. 2017;9:71–7. doi: 10.1016/j.jeoa.2016.08.002.

8. Temple JB, Rice JM, McDonald PF. Mature age labour force participation and the life cycle deficit in Australia: 1981–82 to 2009–10. The Journal of the Economics of Ageing. 2017;10:21–33. doi: 10.1016/j.jeoa.2017.08.001.

9. Minami U, Nishi M, Fukaya T, Hasebe M, Nonaka K, Koike T, et al. Effects of the Change in Working Status on the Health of Older People in Japan. PLoS One. 2015;10(12):e0144069. Epub 2015/12/04. doi: 10.1371/journal.pone.0144069. PubMed PMID: 26633033; PubMed Central PMCID: PMCPMC4669179.

10. ILO. World social protection report 2017–19: Universal social protection to achieve the Sustainable Development Goals. 2017.

11. Adhikari R, Soonthorndhada K, Haseen F. Labor force participation in later life: evidence from a cross-sectional study in Thailand. BMC Geriatr. 2011;11:15. Epub 2011/04/12. doi: 10.1186/1471-2318-11-15. PubMed PMID: 21477283; PubMed Central PMCID: PMCPMC3080298.

12. Dantas RG, Perracini MR, Guerra RO, Ferriolli E, Dias RC, Padula RS. What are the sociodemographic and health determinants for older adults continue to participate in work? Arch Gerontol Geriatr. 2017;71:136–41. Epub 2017/05/02. doi: 10.1016/j.archger.2017.04.005. PubMed PMID: 28458105.

13. Chowdhury P, Mohanty I, Singh A, Niyonsenga T. Informal sector employment and the health outcomes of older workers in India. PLOS ONE. 2023;18(2):e0266576. doi: 10.1371/journal.pone.0266576.

14. Dhar A. Workforce participation among the elderly in India: Struggling for economic security. The Indian Journal of Labour Economics. 2014;57(3):221–45.

15. United Nations. World Population Ageing 2015. D. o. E., & Social Affairs, P. D. United Nations New York, NY. 2015.

16. Reddy AB. Labour force participation of elderly in India: patterns and determinants. International Journal of Social Economics. 2016;43(5):502–16. doi: 10.1108/ijse-11-2014-0221.

17. Schulte PA, Grosch J, Scholl JC, Tamers SL. Framework for Considering Productive Aging and Work. Journal of Occupational and Environmental Medicine. 2018;60(5).

18. Visaria A, Dommaraju P. Productive aging in India. Soc Sci Med. 2019;229:14–21. Epub 2018/08/02. doi: 10.1016/j.socscimed.2018.07.029. PubMed PMID: 30064707.

19. Teerawichitchainan B, Prachuabmoh V, Knodel J. Productive aging in developing Southeast Asia: Comparative analyses between Myanmar, Vietnam and Thailand. Soc Sci Med. 2019;229:161–71. Epub 2018/10/18. doi: 10.1016/j.socscimed.2018.09.053. PubMed PMID: 30327161.

20. Dogra S, Dunstan DW, Sugiyama T, Stathi A, Gardiner PA, Owen N. Active Aging and Public Health: Evidence, Implications, and Opportunities. Annual Review of Public Health. 2022;43(Volume 43, 2022):439–59. doi: 10.1146/annurev-publhealth-052620-091107.

21. Rajan SI. Demographic ageing and employment in India. Bangkok: International Labour Organization, Regional Office for Asia and the Pacific(ILO Asia-Pacific Working paper series). 2010.

22. Mishra S. Social Security for Unorganised Workers in India. Journal of Social Sciences. 2017;53(2):73–80. doi: 10.1080/09718923.2017.1340114.

23. United Nations. World Population Prospects 2019, Department of Economic and Social Affairs, Population Division 2019 [cited 23 September 2021]. Available from: https://population.un.org/wpp/Graphs/DemographicProfiles/Line/356.

24. Anand A. Inpatient and outpatient health care utilization and expenditures among older adults aged 50 years and above in India. Health Prospect. 2016;15(2):11–9. doi: 10.3126/hprospect.v15i2.15831.

25. Yadav S, Arokiasamy P. Understanding epidemiological transition in India. Global Health Action. 2014;7(1):23248. doi: 10.3402/gha.v7.23248.

26. Chowdhury P, Garg MK, Ladusingh L. Morbidity Pattern of Elderly in India. Issues on Health and Healthcare in India: Springer; 2018. p. 445–58.

27. Mini GK, Thankappan KR. Pattern, correlates and implications of non-communicable disease multimorbidity among older adults in selected Indian states: a cross-sectional study. BMJ Open. 2017;7(3):e013529. Epub 2017/03/10. doi: 10.1136/bmjopen-2016-013529. PubMed PMID: 28274966; PubMed Central PMCID: PMCPMC5353268.

28. Puri P, Singh SK. Patterns and predictors of non-communicable disease multimorbidity among older adults in India: evidence from longitudinal ageing study in India (LASI), 2017– 2018. Journal of Public Health Policy. 2022;43(1):109–28. doi: 10.1057/s41271-021-00321-x.

29. Prinja S, Singh MP, Aggarwal V, Rajsekar K, Gedam P, Goyal A, et al. Impact of India’s publicly financed health insurance scheme on public sector district hospitals: a health financing perspective. The Lancet Regional Health - Southeast Asia. 2023;9. doi: 10.1016/j.lansea.2022.100123.

30. Hooda SK. Out-of-pocket Payments for Healthcare in India: Who Have Affected the Most and Why? Journal of Health Management. 2017;19(1):1–15. doi: 10.1177/0972063416682535.

31. Selvaraj S, Farooqui HH, Karan A. Quantifying the financial burden of households’ out-of-pocket payments on medicines in India: a repeated cross-sectional analysis of National Sample Survey data, 1994-2014. BMJ Open. 2018;8(5):e018020. Epub 20180531. doi: 10.1136/bmjopen-2017-018020. PubMed PMID: 29858403; PubMed Central PMCID: PMCPMC5988077.

32. Agrawal G, Mishra A. Universal Health Coverage Initiatives for Elderly-A Review of Ayushman Bharat Program in India. Aging Medicine and Healthcare. 2021;12:34–40.

33. India H. State of elderly in India. New Delhi, available at: https://www.helpageindia.org/images/pdf/state-elderly-india-2014.pdf (Accessed 18 April 2023). 2014.

34. Jeyashree K, Suliankatchi Abdulkader R, Kathirvel S, Chinnakali P, Kumar Mv A. Profile of and expenditure on morbidity and hospitalizations among elderly-Analysis of a nationally representative sample survey in India. Arch Gerontol Geriatr. 2018;74:55–61. Epub 2017/10/11. doi: 10.1016/j.archger.2017.09.007. PubMed PMID: 28992514.

35. Sahoo H, Govil D, James KS, Prasad RD. Health issues, health care utilization and health care expenditure among elderly in India: Thematic review of literature. Aging and Health Research. 2021;1(2):100012. doi: 10.1016/j.ahr.2021.100012.

36. Bharati K, Singh C. Ageing in India: need for a comprehensive policy. J IIM Bangalore Research Paper. 2013;(421).

37. Dhillon P, Ladusingh L. Working life gain from gain in old age life expectancy in India. Demographic Research. 2013;28:733–62. doi: 10.4054/DemRes.2013.28.26.

38. Gokhale SD. Towards a Policy for Aging in India. Journal of Aging & Social Policy. 2003;15(2-3):213–34. doi: 10.1300/J031v15n02_13.

39. Goli S, Reddy AB, James K, Srinivasan V. Economic independence and social security among India’s elderly. Econ Polit Wkly. 2019;54(39):32–41.

40. Sarvesetty C. An Analysis of The Ayushman Bharat Pradhan Mantri Jan Aarogya Yojana (PMJAY) Policy with Respect to the Various Theories of Development with a Special Focus on the Marxist Ideology. Available at SSRN 4528821. 2023.

41. Chowdhury P, Singh A. Are Informal Older Workers Utilizing Less Healthcare Services? Evidence from the Longitudinal Ageing Study in India, Wave-1. Journal of Population Ageing. 2024. doi: 10.1007/s12062-024-09458-5.

42. Census of India. Census of India : Census tables B series 2011 [8 June 2020]. Available from: https://censusindia.gov.in/census.website/data/census-tables.

43. Vives A, Gray N, Gonzalez F, Molina A. Gender and Ageing at Work in Chile: Employment, Working Conditions, Work-Life Balance and Health of Men and Women in an Ageing Workforce. Ann Work Expo Health. 2018;62(4):475–89. Epub 2018/03/27. doi: 10.1093/annweh/wxy021. PubMed PMID: 29579144.

44. Tong Y, Chen F, Su W. Living arrangements and older People’s labor force participation in Hong Kong, 1986-2016. Soc Sci Med. 2019;229:50–9. Epub 2018/10/28. doi: 10.1016/j.socscimed.2018.10.011. PubMed PMID: 30366593.

45. Giles J, Wang D, Cai W. The labor supply and retirement behavior of China’s older workers and elderly in comparative perspective: The World Bank; 2011.

46. Luo Y, Pan X, Zhang Z. Productive activities and cognitive decline among older adults in China: Evidence from the China Health and Retirement Longitudinal Study. Soc Sci Med. 2019;229:96–105. Epub 2018/10/03. doi: 10.1016/j.socscimed.2018.09.052. PubMed PMID: 30274688.

47. Tomioka K, Kurumatani N, Hosoi H. Beneficial effects of working later in life on the health of community-dwelling older adults. Geriatr Gerontol Int. 2018;18(2):308–14. Epub 2017/11/03. doi: 10.1111/ggi.13184. PubMed PMID: 29094489.

48. Buckley J, Tucker G, Hugo G, Wittert G, Adams RJ, Wilson DH. The Australian baby boomer population--factors influencing changes to health-related quality of life over time. J Aging Health. 2013;25(1):29–55. Epub 2012/11/13. doi: 10.1177/0898264312464885. PubMed PMID: 23142925.

49. Forbes MK, Spence KM, Wuthrich VM, Rapee RM. Mental Health and Wellbeing of Older Workers in Australia. Work, Aging and Retirement. 2015;1(2):202–13. doi: 10.1093/workar/wav004.

50. Silver MP, Dass AR, Laporte A. The effect of post-retirement employment on health. The Journal of the Economics of Ageing. 2018. doi: 10.1016/j.jeoa.2018.11.005.

51. Jang S-N, Cho S-I, Chang J, Boo K, Shin H-G, Lee H, et al. Employment status and depressive symptoms in Koreans: results from a baseline survey of the Korean Longitudinal Study of Aging. 2009;64(5):677–83.

52. IIPS., NPHCE., MoHFW., HSPH., USC. Longitudinal Ageing Study in India (LASI) Wave 1, 2017-18, India Report. Mumbai: International Institute for Population Sciences, 2020.

53. NUEPA. EDUCATIONAL DEVELOPMENT INDEX. Department of Educational Management Information System National University of Educational Planning and Administration. 2009.

54. Cabral J, Vidaurre D, Marques P, Magalhães R, Silva Moreira P, Miguel Soares J, et al. Cognitive performance in healthy older adults relates to spontaneous switching between states of functional connectivity during rest. Scientific Reports. 2017;7(1):5135. doi: 10.1038/s41598-017-05425-7.

55. Singh P, Govil D, Kumar V, Kumar J. Cognitive Impairment and Quality of Life among Elderly in India. Applied Research in Quality of Life. 2017;12(4):963–79. doi: 10.1007/s11482-016-9499-y.

56. Kumar S, Nakulan A, Thoppil SP, Parassery RP, Kunnukattil SS. Screening for depression among community-dwelling elders: usefulness of the center for epidemiologic studies depression scale. Indian journal of psychological medicine. 2016;38(5):483–5.

57. Peltzer P, Peltzer K. Utilization of complementary and traditional medicine practitioners among older adults in India: results of a national survey in 2016-2017. 2021.

58. Ansari S, Anand A, Hossain B. Multimorbidity and depression among older adults in India: Mediating role of functional and behavioural health. PLOS ONE. 2022;17(6):e0269646. doi: 10.1371/journal.pone.0269646.

59. Austin PC. An Introduction to Propensity Score Methods for Reducing the Effects of Confounding in Observational Studies. Multivariate Behavioral Research. 2011;46(3):399–424. doi: 10.1080/00273171.2011.568786.

60. Li M. Using the Propensity Score Method to Estimate Causal Effects: A Review and Practical Guide. Organizational Research Methods. 2012;16(2):188–226. doi: 10.1177/1094428112447816.

61. Guo S, Fraser MW. Propensity score analysis: Statistical methods and applications: SAGE publications; 2014.

62. Caliendo M, Kopeinig S. SOME PRACTICAL GUIDANCE FOR THE IMPLEMENTATION OF PROPENSITY SCORE MATCHING. Journal of Economic Surveys. 2008;22(1):31–72. doi: 10.1111/j.1467-6419.2007.00527.x.

63. Rosenbaum PR, Rubin DB. The central role of the propensity score in observational studies for causal effects. Biometrika. 1983;70(1):41–55. doi: 10.1093/biomet/70.1.41.

64. Morgan CJ. Reducing bias using propensity score matching. Journal of Nuclear Cardiology. 2018;25(2):404–6. doi: 10.1007/s12350-017-1012-y.

65. StataCorp. Stata Statistical Software: Release 14. doi: 10.1186/s12970-017-0182-y. 2015.

66. Li Y, Xu L, Chi I, Guo P. Participation in productive activities and health outcomes among older adults in urban China. Gerontologist. 2014;54(5):784–96. Epub 2013/09/14. doi: 10.1093/geront/gnt106. PubMed PMID: 24030035.

67. Baxter S, Blank L, Cantrell A, Goyder E. Is working in later life good for your health? A systematic review of health outcomes resulting from extended working lives. BMC Public Health. 2021;21(1):1356. doi: 10.1186/s12889-021-11423-2.

68. Fisher GG, Chaffee DS, Tetrick LE, Davalos DB, Potter GG. Cognitive functioning, aging, and work: A review and recommendations for research and practice. Journal of Occupational Health Psychology. 2017;22(3):314–36. doi: 10.1037/ocp0000086.

69. Dandona L, Dandona R, Kumar GA, Shukla DK, Paul VK, Balakrishnan K, et al. Nations within a nation: variations in epidemiological transition across the states of India, 1990–2016 in the Global Burden of Disease Study. The Lancet. 2017;390(10111):2437–60. doi: 10.1016/S0140-6736(17)32804-0.

70. Khan GA. A critical analysis of vulnerability in informal sector employment in India: Protective mechanisms and adequacy of protection. International Social Science Journal. 2021;71(241-242):197–215. doi: 10.1111/issj.12288.

71. Karan A, Yip W, Mahal A. Extending health insurance to the poor in India: An impact evaluation of Rashtriya Swasthya Bima Yojana on out of pocket spending for healthcare. Social Science & Medicine. 2017;181:83–92. doi: 10.1016/j.socscimed.2017.03.053.

72. Garg S, Bebarta KK, Tripathi N. Performance of India’s national publicly funded health insurance scheme, Pradhan Mantri Jan Arogaya Yojana (PMJAY), in improving access and financial protection for hospital care: findings from household surveys in Chhattisgarh state. BMC Public Health. 2020;20(1):949. doi: 10.1186/s12889-020-09107-4.

73. Garg S, Chowdhury S, Sundararaman T. Utilisation and financial protection for hospital care under publicly funded health insurance in three states in Southern India. BMC Health Services Research. 2019;19(1):1004. doi: 10.1186/s12913-019-4849-8.

74. Narayana D. Review of the Rashtriya Swasthya Bima Yojana. Economic and Political Weekly. 2010;45(29):13–8.

75. Srivastava S, Bertone MP, Parmar D, Walsh C, De Allegri M. The genesis of the PM-JAY health insurance scheme in India: technical and political elements influencing a national reform towards universal health coverage. Health Policy and Planning. 2023;38(7):862–75. doi: 10.1093/heapol/czad045.

76. Yen H-Y, Chi M-J, Huang H-Y. Social engagement for mental health: An international survey of older populations. International Nursing Review. 2022;69(3):359–68. doi: 10.1111/inr.12737.

77. Newman MG, Zainal NH. The value of maintaining social connections for mental health in older people. The Lancet Public Health. 2020;5(1):e12–e3. doi: 10.1016/S2468-2667(19)30253-1.

78. Mackenzie CS, and Abdulrazaq S. Social engagement mediates the relationship between participation in social activities and psychological distress among older adults. Aging & Mental Health. 2021;25(2):299–305. doi: 10.1080/13607863.2019.1697200.

